## Supplemental Data and Methods for "Genome Sequencing is Critical for Forecasting Outcomes following Congenital Cardiac Surgery"

Authors: see main text for authors and affiliations

October 10, 2024

### Supplemental Data

#### Probabilistic classification of patients

To better define the genetic contribution to severe congenital heart defects (CHD), patients with critical CHDs from the Pediatric Cardiac Genomics Consortium (PCGC) were grouped into five phenotypic categories that reflect underlying structural growth patterns observed during normal human heart development. These five categories are: atrial ventricular canal defects (AVC), conotruncal defects (CTD), laterality defects and heterotaxy (HTX), left ventricular outflow tract defects (LVO), and other cardiac defects (OTH). Grouping patients with similar cardiac structural defects increases the power to detect associations between genetic mutations and cardiac defects and facilitates new hypothesis testing. Classification of patients with CHD defects into unique developmental pathways requires detailed phenotype information.

The Fyler medical coding system provides over 3,000 terms for the granular description of heart malformations. Combinations of the Fyler terms can be used to assign patients to the five CHD categories described above, but manual medical record review is time consuming and typically requires expert knowledge of CHD. To expedite patient classification, we used machine learning techniques to train an extreme gradient-boosted decision tree method to automatically classify CHD patients into the five CHD categories.

We trained and implemented an extreme gradient boosted decision classifier using XGBoost implemented in a user-friendly front-end, XGBoost.jl, available from the Distributed (Deep) Machine Learning Community. This machine-learning method applies supervised learning to classification problems for large data sets with many variables. The method requires training data with accurately known labels (*i.e.,* a truth set) to first identify variables to be used as informative features. A classifier is then created that uses those informative features in optimized decision trees. Binary or probabilistic classifiers can be created. To classify a patient into one of the five CHD categories, we implemented a probabilistic classifier.

A boosted gradient classifier was trained using 3,000 CHD patients from the PCGC. All patient phenotype data was obtained from the HeartsMart database. Phenotype data consisted of 698 phenotype features (Fyler terms) recoded as binary traits (see Supplementary Table 8). Each individual was first manually assigned to one of the five cardiac categories by a trained cardiologist to create a training data set. A five-fold cross-validation approach was then used to train the classifier. We minimized the multiclass log-loss parameter and its standard deviation while simultaneously minimizing the overall classification error. A final model was optimized at a maximum tree depth of 16 and learning rate of 0.15. The final trained classifier model is available as a binary file that can be used with XGBoost libraries. The learning landscape for model optimization is shown in Supplemental Figure 1.

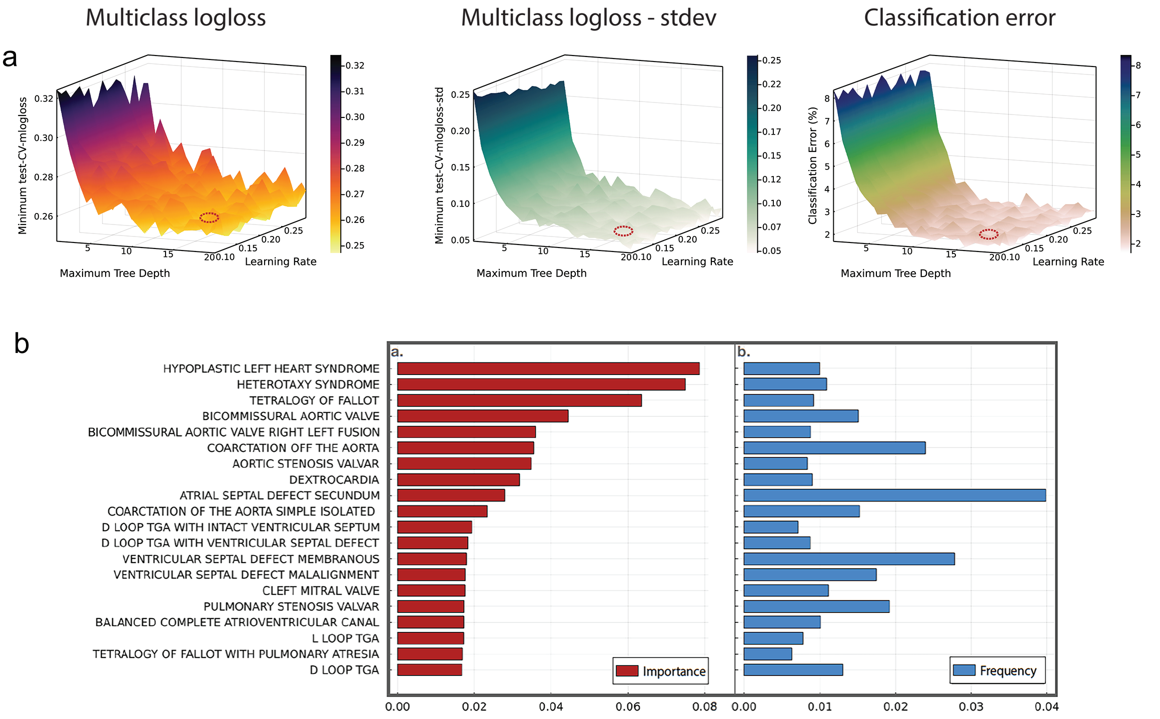


**Supplemental Figure 1. Training landscape for the CHD phenotype classifier. (a)** Panel 1) The multiclass cross-validated loss metric (mlogloss) across all training runs is plotted as a function of the maximum tree depth and model learning rate. The loss function is minimized at a tree depth of 16 and a learning rate of 0.15. Panel 2) The corresponding training landscape for the standard deviation of the mlogloss values shown in Panel 1. Panel 3) The cross-validation training error rate is stable at tree depths of more than ten and remains low as the scoring metric (mlogloss) and its standard deviation approach their optimal (minimal) values. The red ellipses indicate the approximate locations of the optimized values used in the final classifier. **(b)** Importance and usage frequency are shown for the top twenty machine-learning Fyler descriptors used to classify CHD patients into five phenotype categories. The first panel (a) shows the relative contribution of the top 20 features to the final classifier model across all decision trees. Importance (feature gain) represents the learned utility of that feature for decision tree construction. The terms hypoplastic left heart syndrome, heterotaxy syndrome, and tetralogy of Fallot have the highest utility and map patients with those Fyler terms almost exclusively to the LVO, HTX, and CTD phenotype categories, respectively. The second panel indicates the relative usage frequency of the features shown in the first panel used to construct decision trees in the final classifier model.

We provide an example of how individuals are assigned to a phenotype class (Supplemental Figure 2). The prediction probabilities emitted by the classifier sum to one across the five class categories. The predicted CHD phenotype class for a patient is chosen as the phenotype class with the highest probability. Using the trained XGBoost classifer and a set of Fyler descriptions, every available PCGC CHD patient (14,765) was assigned to one of the five CHD phenotype classes (see Supplemental Table 11).

**Prediction probabilities for five patients in the cross-validated training set**

| | AVC | CTD | HTX | LVO | OTH | Predicted class | True class[1] |
|---|---|---|---|---|---|---|---|
| Patient 1 | 0.00007 | 0.99035 | 0.00041 | 0.00582 | 0.00336 | CTD | CTD |
| Patient 2 | 0.00004 | 0.00725 | 0.00061 | 0.99196 | 0.00014 | LVO | LVO |
| Patient 3 | 0.00195 | 0.90099 | 0.00629 | 0.02659 | 0.06418 | CTD | CTD |
| Patient 4 | 0.00304 | 0.20468 | 0.02307 | 0.26297 | 0.50624 | OTH | LVO |
| Patient 5 | 0.00438 | 0.03771 | 0.88106 | 0.00049 | 0.07635 | HTX | HTX |

**Supplemental Figure 2. Probabilistic classification of CHD patients.** Prediction probabilities for the five CHD phenotype classes (AVC, CTD, HTX, LVO, OTH) are shown for five PCGC CHD patients. The predicted class is the one with the highest probability. The true class is the manually-assigned published phenotype for that individual. In four of the five patients, the highest predicted probability (green) matches the true class. In these cases, the classifier accurately predictes the patients' CHD phenotypes. For patient four, the predicted class is OTH (red), but the true class is LVO. This prediction is counted as an error and is a false negative for LVO and false positive for OTH. The quality of the trained model can be evaluated by quantifying the correct and incorrect predictions in a confusion matrix (see Supplemental Table 9; [1]PMID:28991257)

To further assess the quality of the automated classification, we calculated the prediction differential for each patient. This metric is defined as the best prediction probability minus the next best prediction probability times 100. This value ranges, theoretically, from zero (completely ambiguous) to 100 (unambiguous), with the general expectation that the higher the score, the greater the confidence in the prediction. The expectation is valid for a well-optimized model.

The distribution of prediction differentials for the 2,932 correctly assigned patients is dominated by higher scores (Supplemental Figure 3a) indicating high-confidence predictions. The distribution of the 68 misclassified samples is instructive with regard to classifier accuracy. This distribution is dominated by low scores, as expected (Supplemental Figure 3b). There are, however, a very small number of misclassified samples with scores > 50, suggesting that rare combinations of certain Flyer descriptors are not fully accounted for by the model. These rare events occurred in < 0.7% of all patient classifications. The prediction differentials can also serve as a quality-guide, with low values indicating assignments that should be evaluated by an experienced cardiologist. Additionally, the distribution of prediction differentials should be carefully evaluated if the classifer is used in other geographically or ethnically diverse CHD cohorts.

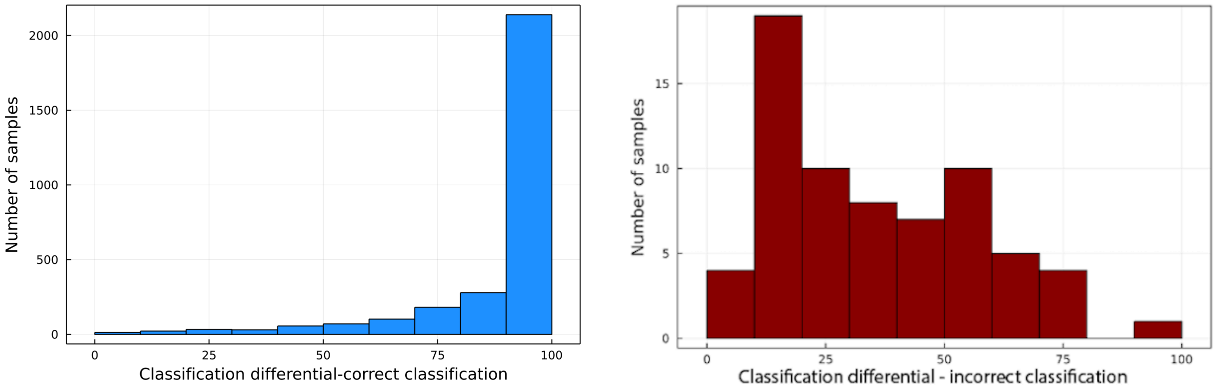


**(a)** Classification differentials for 2,932 correctly classified samples. The majority of correctly classified samples have a differential score of at least 75 (mean = 89).

**(b)** Classification differentials for 68 incorrectly classified samples. The majority of incorrectly classified samples have a differential score less than 50 (mean = 34).

**Supplemental Figure 3. Classification differentials for 3,000 training samples.** The classification differentials, the difference between the highest classification probability and the next best classification probability times 100 for each patient, are exponentially distributed and inversely related. Note: the y-axis scale for the number of samples differs between panels (a) and (b).

#### Gene lists

Gene lists were obtained using the Reactome pathway browser [reactome.org] and have been previously described [PMID:31624253]. A small number of genes are shared among pathways (Supplemental Figure 4). Chromatin-modifying genes overlap with genes in the non-canonical Wnt pathway. Genes for structural and motile cilia production overlap with genes in the *FOXJ1* pathway, which are involved in motile cilia production. Some signal transduction pathway genes are found in the non-canonical Wnt pathway but are not found in any of the other gene lists. The CHD gene list represents genes previously associated with CHD but these genes are not pathway specific.

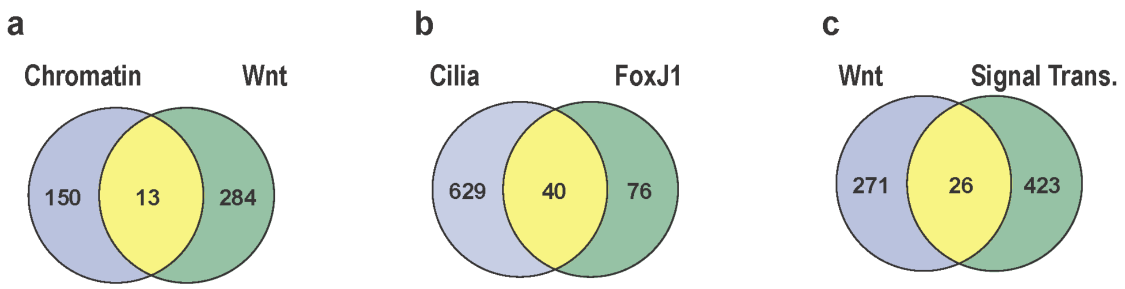


**Supplemental Figure 4. Relationships among gene lists.** Venn diagrams illustrate the overlap among key gene lists used in the main text. a) Chromatin-modifying genes (163) and non-canonical *WNT*-related genes (297), b) Cilia genes and *FOXJ1* related genes, and c) non-canonical *WNT*-related genes (297) and signal transduction genes (449).

#### Bayesian networks and variable correlations

The complex relationships among genetic, phenotypic, and surgical outcomes, combined with a relatively low number of CHD patients across multiple variable domains, make Bayesian networks a natural choice for understanding the factors influencing surgical outcomes. Bayesian networks leverage information from prior observational data for risk estimation and can discover new relationships among variables while also accounting for the effects of covariates and non-linear relationships. Risk outcomes and their confidence intervals can be estimated even when events are rare.

We present Bayesian network models for genetic, phenotypic, and surgical variables in patients with laterality defects (HTX) and left ventricular outflow tract obstructions (LVO), two of the most serious classes of pediatric congenital heart disease. Each model is a directed acyclic graph (DAG) of variables whose relationships were learned using an exact method to systematically find a globally optimal Bayesian network structure [arXiv:1206.6875].

Briefly, the learning algorithm performs the following steps. For each variable, 1) the best parent–offspring pairs are identified ($n2n - 1$ combinations); then 2) the best offspring–parent sets are identified (also $n2n - 1$ combinations). Pairs and sets with the best local score, as determined using the Bayesian Information Criterion (BIC) metric are selected. Minimal sinks are then identified for all parent–offspring sets ($2n$ combinations). These sets are ordered and assembled into networks, where each network is scored as the sum of the local scores for the parents and offspring sets. A best network having the highest score is selected. Exact networks provide an accurate representation of the joint probability distribution for all variables, computational constraints limit these networks to a small number of variables (see Supplemental Table 13 for all network variables).

To choose which surgical variables to include in the networks, each surgical variable was screened for conditional dependence on a CHD phenotype category. Only surgical variables showing a positive association (*i.e.,* increased risk) with a phenotype were retained. We then limited the final set of variables to include potentially actionable surgical variables of high clinical interest (cardiac arrest, prolonged ventilation time, and mortality).

Determining the structure of a Bayesian network can be influenced by correlation among variables. Collinear and multiply collinear variables can mask variable importance and reduce the predictive power of a Bayesian network. We tested the pairwise correlations among variables in the networks presented in the main text and found only low correlations among these variables ($0.11 \leq \rho \leq 0.3$) , except for the expected high correlation between LVO and HLHS ($\rho = 0.59$) (Supplemental Figure 5 a and b). We note that in the Bayesian networks, the assump-tion of non-independence between the parents and non-descendant nodes applies to the learning and structure of the Bayesian network. This assumption does not imply that variables that are conditionally independent in the network cannot be correlated. Conversely, two apparently non-correlated variables are not necessarily independent once they are examined in network context because one or more other network variables may reveal a new conditional dependency.

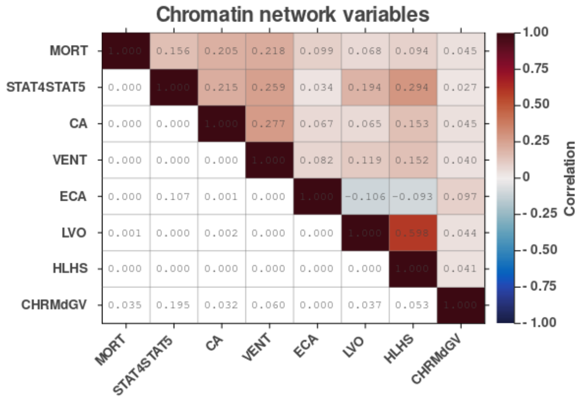


**(a)** Pairwise correlations among variables in the Bayesian network for patients with damaged chromatin-modifying genes, related phenotypes, and surgical outcomes (see Main text – Figure 1a).

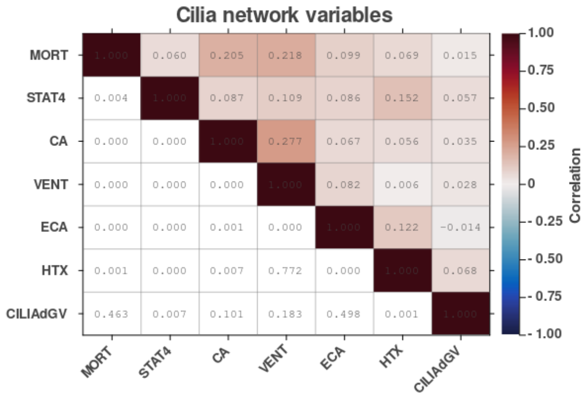


**(b)** Pairwise correlations among variables in the Bayesian network for patients with damaged cilia-related genes, related phenotypes, and surgical outcomes (see Main text – Figure 1b).

**Supplemental Figure 5. Spearman correlations and P-values among variables used in each Bayesian network as presented in the main text.** In both panels, correlations are shown in the upper triangle and diagonal. P-values are shown in the lower triangle. Correlations among variables are low ($\rho < 0.3$) and often significant (Bonferroni corrected P-values: P < 0.0017 and P < 0.0023, respectively). No correlations are collinear or multiply collinear using a standard cutoff of $\rho \leq 0.7$, indicating that correlations among variables is not expected to have a substantial impact on network structure discovery.

The accuracy of network inferred risk estimates can be affected by sample size. Many studies advocate Bayesian analysis for low sample size applications. With low sample sizes, accurate posterior probabilities and risk estimates are highly dependent on informative and unbiased prior probability estimates [doi.org/10.1080/10705511.2016.1186549]. Our initial probability estimates for variables used in the study are based on 2,253 CHD patients from multiple surgical sites and are, therefore, likely to be representative probability estimates for a critical CHD surgical cohort and without strong biases. Observed variable frequencies are listed in Supplemental Table 12.
